# Hypophosphataemia in suspected seizures evaluated in the first seizure clinic and neurology consults

**DOI:** 10.1101/2025.06.26.25329859

**Authors:** SNM Binks, D Zorkin, B Liem, A Sen

**Author notes:** Corresponding author to whom correspondence should be addressed: Prof Arjune Sen. **Ethics/patient consent statement** The audit was registered with Oxford University Hospitals NHS Foundation Trust; reference number 8576. It was reviewed by Oxford University Hospitals Foundation NHS Trust where it was deemed that further Ethics Approval was not required.

## Abstract

Transient loss of consciousness (TLoC) is a leading cause of referrals to acute neurological services. A witness account is often lacking and ancillary investigations are a critical diagnostic adjunct. Hypophosphataemia was recently identified as a potential maker of epileptic seizures in ward and emergency department presentations.

We evaluated the real-world utility of checking phosphate in an unselected cohort of people presenting with TLoC. We retrospectively reviewed 182 episodes (91 referrals to first seizure clinic and 91 consults) from 173 patients. We assessed nine pre-specified serological markers frequently measured in people presenting with TLoC. Raw *P*-values comparing mean levels showed significant difference between epileptic seizures and non-epileptic episodes only for phosphate (0.98 vs. 1.19 mmol/L, *P*=0.006) and lactate (2.82 vs. 1.82 mmol/L, *P*=0.007). No blood biomarkers were significant after multiple comparison correction, although a phosphate below 0.8mmol/L was significantly more likely to associate with epileptic seizures than non-epileptic episodes (17/64, 26.5% vs. 7.4%, 2/27, *P*=0.049). Logistic regression showed that a model including lactate and phosphate was most accurate to predict epileptic seizures with an area-under-the ROC curve of 0.728 (96% CI 0.607-0.848). Checking serum phosphate may be valuable in helping to determine the aetiology of an episode of TLoC.

**Plain language summary:** We studied blood test results of 173 people suspected of having had an epileptic seizure who presented to a UK hospital neurology service. Although blood phosphate tests were infrequently requested, our results suggest a low phosphate level could be useful to help distinguish between epileptic and non-epileptic attacks.

## Introduction

Transient loss of consciousness (TLoC) events, incorporating both suspected seizures and other causes of blackouts, are a leading cause of acute neurological presentations. They account for ∼1% of Emergency Department (ED) visits^1^ and around one-sixth of new referrals to Neurology Outpatient Departments.^2^ In most institutions, daily neurological practice involves multiple assessment types for people presenting with TLOC, including phone consults and telehealth. In these circumstances, precise seizure classification may not be possible although advice on episode management is still required. A witness account is essential in differentiating between different causes of TLoC, yet in the UK-based National Audit of Seizure Management in Hospitals, this was only attempted in approximately 70% of first seizure evaluations at the point of contact.^3^ Ancillary evidence provided by supporting investigations can therefore be a useful diagnostic adjunct.

Phosphate was recently shown to be the most profoundly altered electrolyte after generalised tonic-clonic seizures.^4^ Hypophosphataemia has since emerged as a potential biomarker of epileptic seizures compared to other causes of TLoC in European EDs.^5–7^ Low phosphate has also been detected in other mammalian species, for example dogs presenting with seizures.^8^ The frequency with which phosphate levels are checked for people presenting with TLoC in routine clinical care remains to be clarified.

We audited the frequency with which phosphate was measured in TLoC presentations in a real-world UK setting. Our goal was to understand the utility of post-ictal phosphate levels in accurately differentiating epileptic seizures from non-epileptic episodes, including functional dissociative seizures (FDS), in undifferentiated patients.

## Methods

We retrospectively identified cases that were referred to either a first seizure clinic (2021-2023) or to hospital-based on-call neurological services (February-June 2023). Clinical variables captured were age, sex, and time to blood test. Recorded serological variables consisted of nine pre-specified markers in: haematology (neutrophil count); electrolytes (sodium, potassium, calcium, magnesium and phosphate); infection (C-reactive protein (CRP)) and metabolism (glucose and lactate).

Each episode was rated as an epileptic seizure or non-epileptic (including FDS) through detailed notes review. Where a final disposition was not reached, two authors (SNB, AS) reviewed the entire case file to best determine whether the event was epileptic in aetiology or not. This was done independent of serological findings.

Statistics were performed in R (v4.0.3 and v4.4.0). We compared between-group differences for demographic and biochemical features in the epileptic seizure and non-epileptic groups with binomial tests (chi squared or Fisher’s exact test for categorical variables, t-test or Wilcoxon test for continuous variables). Fisher’s test was used if any group compared five or fewer individuals. T-test was used for normally, and Wilcoxon test for non-normally, distributed data, with normality assessed via the Shapiro-Wilk test. Correction for multiple comparisons was performed with Holm’s method. We used Spearman’s test for correlation. Logistic regression was carried out in base R, stepwise regression using the MASS function, and area under the curve (AUC) with the pROC function. Data visualisation was handled with ggplot2 and pROC. Missing data were excluded from relevant analyses. Significance was set at two-sided *P*<0.05.

The audit was registered with Oxford University Hospitals NHS Foundation Trust; reference number 8576.

## Results

Altogether, 182 episodes (91 each first seizure clinic /liaison consultation), were analysed. Overall, 126 (69%) were epileptic seizures. These events derived from 173 patients (78 women, mean age 51 years). Full demographic details are in Table 1. The proportion of requested blood tests across the nine pre-specified parameters ranged from 176/182 (97%) for sodium to 91/182 (50%) for phosphate (Figure 1A).

**Table 1.**
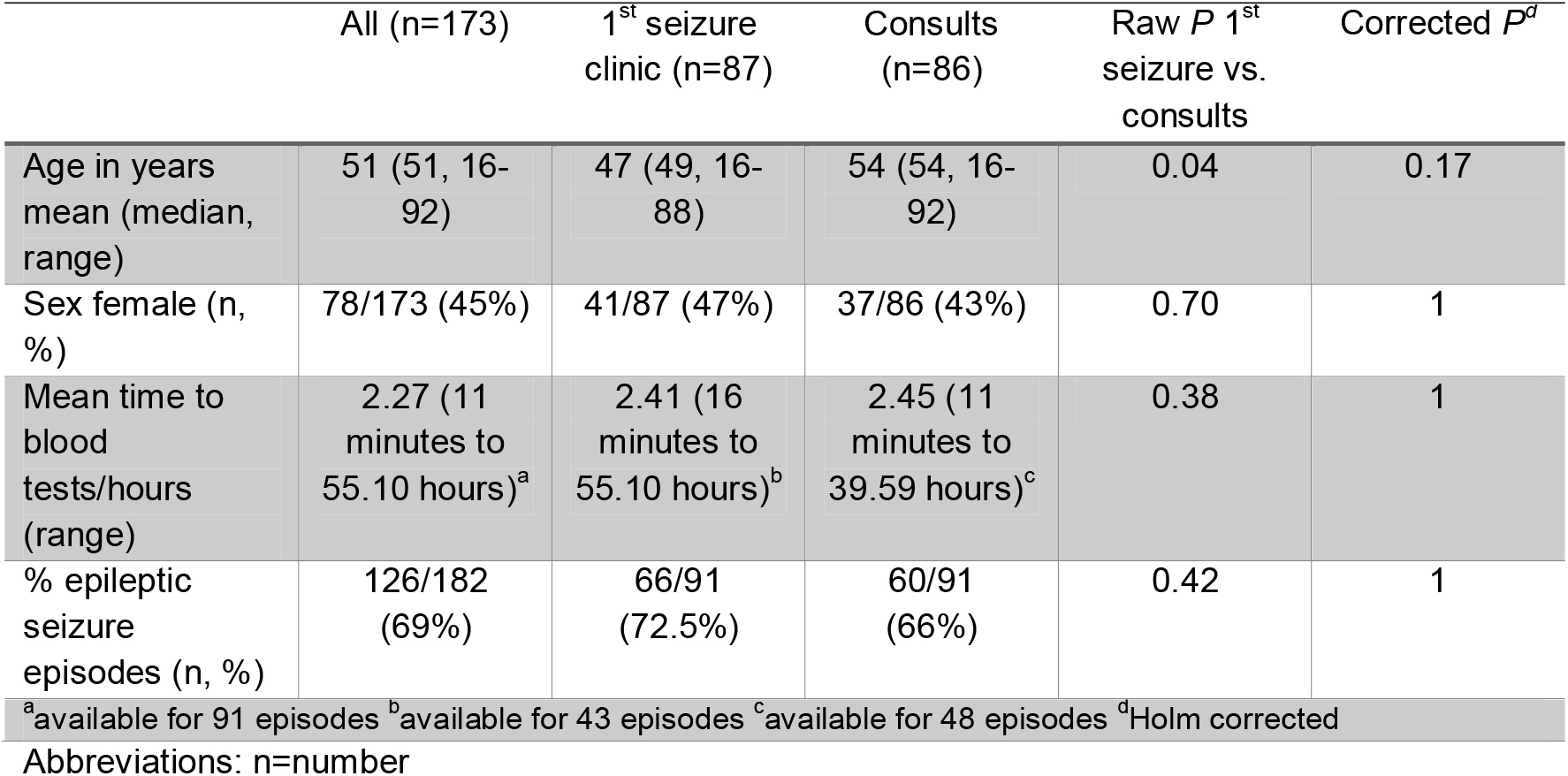
Key demographics by total cohort, and by patients referred to first seizure clinic or consults service.

**Figure 1.**
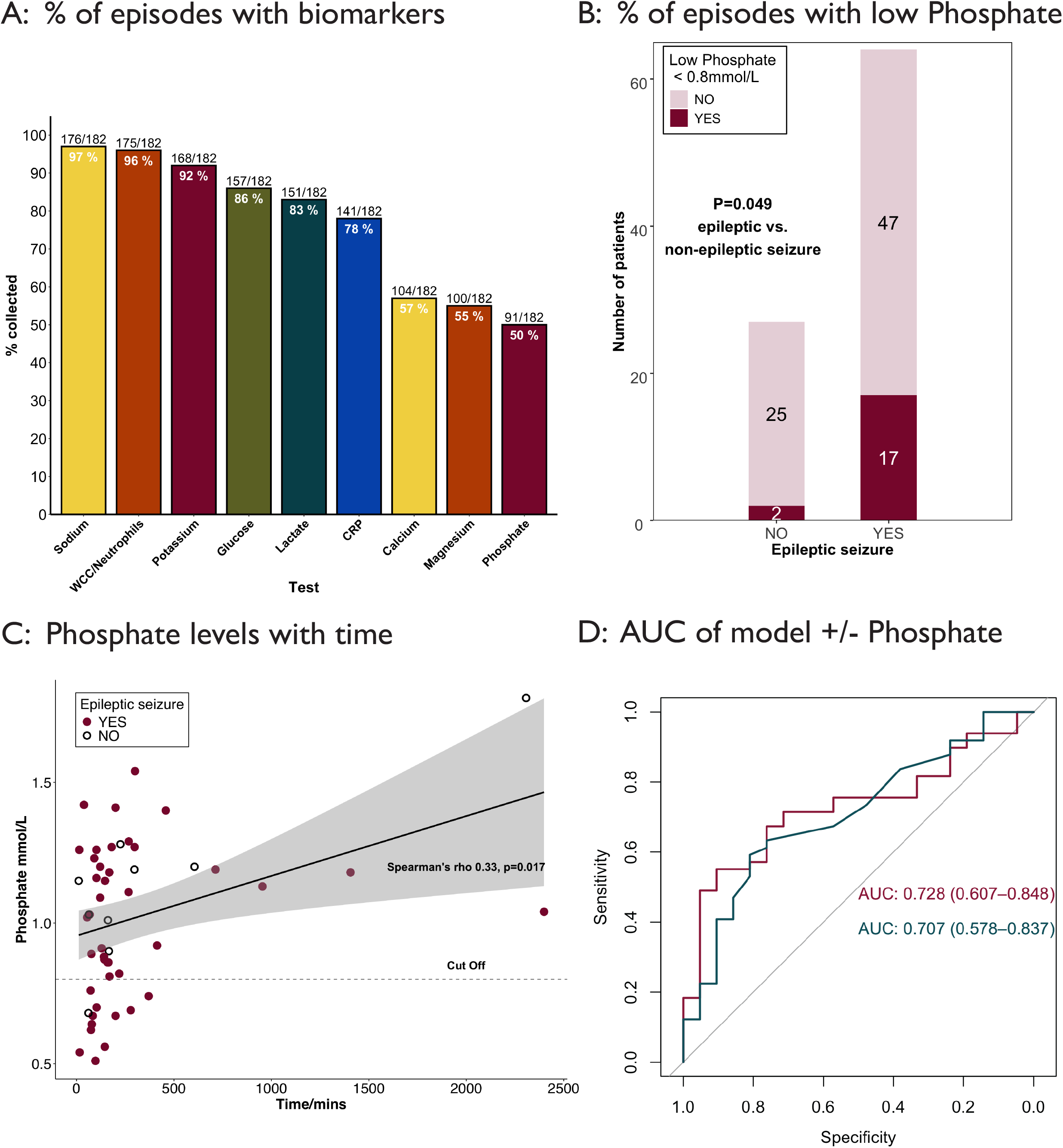
**[A]** Bar chart showing proportion of episodes in which the pre-specified variables were collected, plotted in descending order of frequency. Numbers at the top of the bar give proportion of episodes in which each parameter was collected, and those within the bar represent the percentage. **[B]** Stacked bar chart. A phosphate <0.8 mmol/L was present in 26.5% (17/64) epileptic seizures and 7.4% (2/27) non-epileptic episodes (*P*=0.049) (Fisher’s exact test; dark red shows low phosphate and pale red shows phosphate in normal range). **[C]** Scatter plot of phosphate levels (y axis) against time from episode onset (x axis). Values below the threshold (<0.8mmol/L) were only detected at a maximum of 6.10 hours. Dark red dots represent seizure cases and black dots, non-epileptic seizure cases. The dotted line marks the cut-off for a low phosphate level in our institution (<0.8mmol/L). Shaded area = standard error. **[D]** AUC of a logistic regression model including only lactate level (blue line) or lactate with phosphate level (dark red line). AUC values with 95% confidence intervals are depicted on the graph. **Abbreviations:** AUC, area under the curve; CRP, C-reactive protein; WCC, white cell count.

Per-group biochemical parameters are shown in Table 2. Raw *P*-values comparing mean levels showed significant difference between epileptic seizure and non-epileptic event only for phosphate (0.98 vs. 1.19 mmol/L; *P*=0.006) and lactate (2.82 vs. 1.82 mmol/L; *P*=0.007). No blood biomarkers were significant after multiple comparison correction, but a phosphate below the lower limit of normal in our institution (<0.8mmol/L) was significantly more likely in an epileptic seizure than non-epileptic episodes (17/64, 26.5% vs. 2/27, 7.4%, *P*=0.049) (Figure 1B). There was a modest, but significant, correlation between phosphate level and time from blood draw. All phosphate levels below the threshold for the lower limit of normal were taken within 6.10 hours from episode onset (Figure 1C). Taken together, these findings are consistent with a post-ictal decrease in phosphate being detectable at up to six hours post-ictally.

**Table 2.** Per-group biochemical parameters.

| Parameter – mean (median, range) | All (n=182 episodes) | Epileptic Seizure (n=126 episodes) | Non-epileptic (n=56 episodes) | Raw *P* seizure vs. non-epileptic episode | Corrected *P*<sup>a</sup> |
| --- | --- | --- | --- | --- | --- |
| Sodium (mmol/L) | 138 (139, 121-147) | 138 (139, 121-147) | 138 (139, 126-143) | 0.83 | 1 |
| Neutrophils (x 10<sup>9</sup>/L) | 7.42 (6.52, 1.32-20.82) | 7.48 (6.7, 1.65-19.6) | 7.27 (5.91, 1.32-20.82) | 0.37 | 1 |
| Potassium (mmol/L) | 4.10 (4.10, 2.9-5.5) | 4.05 (4, 2.9-5.2) | 4.18 (4.1, 3.3-5.5) | 0.10 | 0.61 |
| Glucose (mmol/L) | 6.65 (6, 3.3-15.7) | 6.60 (6, 3.8-15.7) | 6.77 (6.1, 3.3-14.3) | 0.78 | 1 |
| Lactate (mmol/L) | 2.53 (1.5, 0.6-19) | 2.82 (1.7, 0.6-19) | 1.82 (1.3, 0.6-11.9) | 0.007 | 0.059 |
| CRP (mg/L) | 15.38 (3.4, 0.2-236.2) | 11 (2.95, 0.2-91.6) | 24.8 (3.9, 0.3-236.2) | 0.32 | 1 |
| Calcium (mmol/L) | 2.35 (2.33, 2.15-2.60) | 2.33 (2.32, 2.15-2.33) | 2.38 (2.36, 2.19-2.60) | 0.03 | 0.23 |
| Magnesium (mmol/L) | 0.82 (0.82, 0.57-1.16) | 0.82 (0.81, 0.57-1.16) | 0.82 (0.82, 0.65-0.94) | 0.94 | 1 |
| Phosphate (mmol/L) | 1.04 (1.10, 0.42-2.13) | 0.98 (0.98, 0.42-1.54) | 1.19 (1.16, 0.66-2.13) | 0.006 | 0.056 |

<sup>a</sup>Holm corrected

Finally, we examined the predictive value of phosphate in differentiating epileptic seizures from non-epileptic events. We evaluated levels of all entities which were nominally significant between the groups (calcium, phosphate, and lactate levels) via a stepwise regression model. The best fit model included phosphate and lactate (Supplementary Information). Although a model with only serum lactate was also significant (Supplementary Information), the combined model had a better AUC of 0.728 (95% CI 0.607-0.848) compared to lactate alone (0.707, 95% CI 0.578-0.837; Figure 1D). A stand-alone logistic regression model with an absolute low phosphate was significant (*P*=0.02) with an odds ratio of 4.5 in predicting an epileptic seizure, although confidence intervals were wide (95% CI 0.97-21.2).

## Discussion

We examined the frequency with which phosphate is incorporated into work-up of TLoC in a real-world setting and its role in differentiating epileptic seizures from non-epileptic events. Our investigation covered both in-person and remote reviews, to better reflect what occurs in acute neurological services.

Despite growing evidence that it is the most altered electrolyte in post-seizure physiology, phosphate was also the most rarely requested in this cohort. The low proportion of episodes in which phosphate was requested in routine practice highlights the potential to improve awareness of its potential utility. It is likely the non-significant adjusted *P*-values for numeric phosphate levels in epileptic seizure compared to non-epileptic events, and wide confidence intervals in a stand-alone logistic regression model, reflect, at least in part, loss of power due to missing data. An absolute phosphate level below 0.8mmol/l (the lower end of the normal range at our institution, although institutional cut-offs differ) was more likely in epileptic compared to non-epileptic episodes. Checking phosphate levels therefore represents a simple, inexpensive and helpful addition to assessment of TLoC events.

Our results, in keeping with former studies, also suggest that checking phosphate levels may be informative up to six hours after ictal onset.^4,5^ The mechanism of hypophosphataemia is not confirmed but potentially includes hormonal or exertional effects.^5,9^

Comparable results were found for lactate, although the addition of phosphate to this more traditional biomarker did modestly increase the ability to discriminate epileptic seizures from other events. This could be of particular value when a witness account is not available or in non-specialist settings. Testing of phosphate in low to middle income countries may, for example, enable primary healthcare workers to better determine the potential cause of an episode of TLoC and streamline initial decision making and facilitate appropriate onward referral.

While most previous studies have focussed on generalised convulsive tonic clonic seizures,^4–6,8^ our report links hypophosphataemia to epileptic seizure episodes more broadly. Hypophosphatemia has recently been reported in connection to autoimmune encephalitis with antibodies to leucine-rich glioma-inactivated 1 encephalitis, a condition that usually presents with frequent focal seizures.^10^ This suggests the mechanism of metabolic impact is not solely driven by muscle contraction and broadens the potential value of measuring phosphate levels in episodes of TLoC.

## Limitations

As with any real world study, there are limitations owing to missing data and that we could not always confirm definitive epileptic seizure activity or the type of seizure. The diagnosis of a seizure, except where concurrent EEG captures an event, remains a clinical diagnosis and should not be ruled in, or out, by a single biomarker.

Our study adds to a growing body of evidence demonstrating that serum phosphate can be a helpful supportive test in evaluating potential seizure episodes. We would propose adding serum phosphate to ED first seizure caresets, for example as part of a battery of tests requested when someone presents with TLOC. Prospective studies should audit the utility of such incorporation in improving diagnostic accuracy.

## Data Availability

Data will be made available on reasonable request.

## Acknowledgements

Dr Binks is funded by an NIHR Clinical Lectureship.

## Notes

**Conflicts of interest** Dr Binks, Dr Zorkin, Dr Liem and Professor Sen have no relevant conflicts of interest.

### Competing Interest Statement

The authors have declared no competing interest.

### Author Declarations

The audit was registered with Oxford University Hospitals NHS Foundation Trust; reference number 8576. It was reviewed by Oxford University Hospitals Foundation NHS Trust where it was deemed that further Ethics Approval was not required. All data were deidentified and only analysed in aggregate

## References

1. Huff JS, Morris DL, Kothari RU, Gibbs MA, THE Emergency Medicine Seizure Study Group (EMSSG)*. Emergency Department Management of Patients with Seizures: A Multicenter Study. Academic Emergency Medicine [Internet]. 2001 Jun 28;8(6):622–8. Available from: https://onlinelibrary.wiley.com/doi/10.1111/j.1553-2712.2001.tb00175.x

2. Fuller G. Neurology. GIRFT Programme National Specialty Report. 2021.

3. National Audit of Seizure Management in Hospitals. Data Analysis and methodology report. 2020.

4. Nass RD, Zur B, Elger CE, Holdenrieder S, Surges R. Acute metabolic effects of tonic-clonic seizures. Epilepsia Open. 2019 Dec 1;4(4):599–608.

5. Barras P, Siclari F, Hügli O, Rossetti AO, Lamy O, Novy J. A potential role of hypophosphatemia for diagnosing convulsive seizures: A case-control study. Epilepsia. 2019;60(8):1580–5.

6. Barbella G, Barras P, Rossetti AO, Novy J. Hypophosphatemia compared to classical biomarkers of tonic clonic seizures. Epilepsy Res. 2020 Jul 1;163.

7. Coutinho MP, Faustino P, Ladeira F, Leitão L. Hypophosphatemia as a possible biomarker for epileptic seizures at the emergency department. Seizure. 2023 Oct 1;111:42–4.

8. Kelmer E, Ohad DG, Shamir MH, Chai O, Lavie S, Sutton GA, et al. The diagnostic utility of hypophosphatemia for differentiating generalized tonic-clonic seizures from syncope in dogs: A case control study. Veterinary Journal. 2023 Jan 1;291.

9. Lo YH, Mok KL. Hypophosphataemia in confused half-marathon runners: A report of two cases. Hong Kong Journal of Emergency Medicine. 2020 Nov 1;27(6):380–3.

10. Gadoth A, Nisnboym Ziv M, Alcalay Y, Zubkov A, Schwartz I, Schwartz D, et al. Electrolyte Imbalance in Anti-LGI1 Encephalitis: It Is Not All in Your Head. Vol. 10, Neurology(R) neuroimmunology & neuroinflammation. NLM (Medline); 2023.

