## Supplementary Information for "Hypophosphataemia in suspected seizures evaluated in the first seizure clinic and neurology consults"

**Stepwise logistic regression model-lactate and phosphate levels as predictor**

step.model_audit <- multivariate_audit_no_NA_step %>% stepAIC(trace = FALSE)

summary (step.model_audit)

Call:

glm(formula = Seizure_yes_no ~ Phos_mmol.L + Lactate_mmol.L,

family = binomial, data = audit_draft3_for_step2)

Coefficients:

Estimate Std. Error z value Pr(>|z|)

(Intercept)	1.6772	1.3178	1.273	0.2031

Phos_mmol.L	-1.6099	1.0844 -1.485	0.1377

Lactate_mmol.L	0.4486	0.2344	1.914	0.0556 .

---

Signif. codes: 0 ‘***’ 0.001 ‘**’ 0.01 ‘*’ 0.05 ‘.’ 0.1 ‘ ’ 1

(Dispersion parameter for binomial family taken to be 1)

Null deviance: 85.521 on 69 degrees of freedom

Residual deviance: 75.596 on 67 degrees of freedom

AIC: 81.596

Number of Fisher Scoring iterations: 5

> exp(coef(step.model_audit))

(Intercept)	Phos_mmol.L Lactate_mmol.L

5.3504416	0.1999018	1.5661832

> > 1-pchisq (85.521-75.596, 69-67)

[1] 0.006995417

**Logistic regression model – absolute low phosphate alone as predictor**

Call:

glm(formula = Seizure_yes_no ~ Phos_low, family = binomial, data = audit_draft3_noNA)

Coefficients:

Estimate Std. Error z value Pr(>|z|)

(Intercept)	0.6313	0.2475	2.550	0.0108 *

Phos_lowYES	1.5088	0.7874	1.916	0.0554 .

---

Signif. codes: 0 ‘***’ 0.001 ‘**’ 0.01 ‘*’ 0.05 ‘.’ 0.1 ‘ ’ 1

(Dispersion parameter for binomial family taken to be 1)

Null deviance: 110.66 on 90 degrees of freedom

Residual deviance: 105.77 on 89 degrees of freedom

AIC: 109.77

Number of Fisher Scoring iterations: 4

> exp(coef(low_phos_audit))

(Intercept) Phos_lowYES

1.880000	4.521277

> 1-pchisq (110.66-105.77, 90-89)

[1] 0.02701269

> ors <- exp(coef(low_phos_audit))[1:2]

> cis <- exp(confint.default(low_phos_audit))[1:2,]

> cbind(ors, cis)

ors	2.5 %	97.5 %

(Intercept) 1.880000 1.1573121 3.053973

Phos_lowYES 4.521277 0.9660477 21.160387
